# Analysis of Healthcare Seeking Behavior Among Patients Visiting Public Primary and Secondary Healthcare Facilities in an Urban Indian District

**DOI:** 10.1101/2022.08.31.22279441

**Authors:** Najiya Fatma, Varun Ramamohan

## Abstract

In this work, we examined healthcare seeking behavior (HSB) of patients visiting public healthcare facilities in an urban context. We conducted a cross-sectional survey across twenty-two primary and secondary public healthcare facilities in the South-west Delhi district in India. The survey was designed to ascertain from patients at these facilities their HSB - i.e., on what basis patients decide the type of healthcare facility to visit, or which type of medical practitioner to consult. From each facility visited, we also collected operational information, such as the average number of patients visiting per day, and the medical services provided at each facility. Based on participant responses, we observed that factors such as wait time, prior experience with care providers, distance from the facility, and also socioeconomic and demographic factors such as annual income, educational qualification, and gender significantly influenced preferences of patients in choosing healthcare facilities. We used binomial and multinomial logistic regression to determine associations between HSB and socioeconomic and demographic attributes of patients at a 0.05 level of significance. Our statistical analyses revealed that patients in the lower income group preferred to seek treatment from public healthcare facilities (OR = 3.51, 95% CI = (1.65, 7.46)) irrespective of the perceived severity of their illness, while patients in the higher income group favored directly consulting specialized doctors (OR = 2.71, 95% CI = (1.34, 5.51)). Other factors such as having more than two children increased probability of seeking care from public facilities. This work contributes to the literature by providing quantitative evidence regarding overall patient HSB, especially at primary and secondary public healthcare facilities, regardless of their presenting illness, and operational information regarding healthcare delivery at these facilities. This work can inform policy designed to improve accessibility and quality of care at public primary and secondary healthcare facilities in India.

## 1. Introduction

### 1.1 Motivation

Healthcare seeking behavior (HSB) involves decisions taken by patients on which facility and/or which type of practitioner – for example, private or public, primary care level or higher level of care facility (e.g., a hospital), or a general physician or a specialist - to visit first upon falling ill (Musinguzi et al. 2018). HSB is considered to be an outcome of the complex interaction between the patient’s illness condition and their socioeconomic and demographic characteristics as well as the quality, availability, and accessibility of healthcare services (Deolia et al. 2020). Healthcare facilities are organized in a hierarchical manner across the globe, including in India, wherein lower level facilities provide treatment for common ailments of mild and moderate severity, while higher level facilities mainly focus on specialized care and illnesses of high severity to ensure equitable access to medical services (Tao & Han, 2021). In a hierarchical system, patients are typically assigned a nearby primary healthcare facility for initial visits, and then referred to higher level facilities as required. However, in practice, a significant proportion of patients often bypass lower level facilities and seek care directly at higher level facilities due to multiple factors such as: (a) the perception that good quality care is provided only at specialized facilities, (b) lack of an effective referral mechanism, (c) lower trust for primary care services, and (d) unbalanced allocation of healthcare resources (Lee et al. 2019).

Establishing primary healthcare facilities as the first point of contact in the event of illness/injury has been emphasized by healthcare administrators in multiple countries (Du et al. 2019). One of the major objectives of the Indian Ministry of Health and Family Welfare involves strengthening and integration of the public healthcare delivery system to provide equitable access to quality healthcare to the Indian population (DGHS, 2022). The tendency to not follow the hierarchy of healthcare delivery by a major proportion of patients places an undue burden on medical resources, especially at higher levels of care in the Indian public healthcare system (Bhola et al. 2008). In this context, two questions arise: (a) on what basis do patients decide to seek care from public primary and secondary healthcare facilities in an urban metropolitan district with a significant number of private alternatives, and (b) what factors are associated with patients visiting primary and secondary care facilities upon first falling ill instead of specialized public or private facilities such as hospitals. In this study, we attempt to answer these questions by administering an appropriately designed survey to patients actually visiting public primary and secondary care facilities in the South-west district of the urban metropolitan city of New Delhi, and relate said HSB to patient socioeconomic and demographic attributes. Note that answering the above questions in a comprehensive manner would involve, in addition to conducting the above survey, also surveying patients visiting (a) private facilities providing primary and secondary care, and (b) tertiary care public and/or private facilities upon first falling ill. Thus, this study is the first step towards answering the above questions.

Previous work involving the study of HSB in India has focused primarily on collecting information of patients either from higher level facilities such as hospitals or from patient localities for specific disease conditions (Chadda et al. 2001; Narang, 2010; Marsh et al. 2020; Thomas et al. 2021). Further, information regarding HSB was primarily collected from patients belonging to a specific age group, gender, or income level in rural or semi-urban areas. Hence, in this work, we administered surveys at multiple primary and secondary public healthcare facilities in an urban district in order to determine the overarching HSB of patients visiting these facilities. Understanding HSB, especially with respect to the public healthcare system in densely populated urban areas, is the first step in improving access to affordable and high-quality healthcare in India, especially given the significantly higher costs incurred in accessing healthcare at privately managed facilities (Rao & Sheffel, 2018).

### 1.2 Literature review

We briefly discuss previous work analyzing patient HSB in both international and Indian contexts. Previous studies primarily considered assessing behavior of patients with respect to diseases such as cancer (Habtu et al. 2018; Mubin et al. 2021), tuberculosis (Kaur et al. 2013; Thomas et al. 2021), oral diseases (Deolia et al. 2020), asthma (Ndarukwa et al. 2020), ante- and prenatal care (Shahabuddin et al. 2017; Matsubara et al. 2019), mental illness (Srivastava et al. 2021), non-communicable diseases (Rasul et al. 2019), diabetes (Low et al. 2016), whiplash injury (Tenenbaum et al. 2017), and HIV (Lafort et al. 2016) among others. HSB was analyzed with respect to patient-related attributes such as gender (Thompson et al. 2016; Kapoor et al. 2019; Rana et al. 2020) or a specific age group (Cho et al. 2008; Shrivastava 2014; Srivastava et al. 2021), rurality/urbanity (Nemet & Bailey, 2000; Herberholz & Phuntsho, 2018; Liu et al. 2018; Liu et al. 2019), marital status (Shahabuddin et al. 2017), and socioeconomic status (Matsubara et al. 2019). Socioeconomic attributes such as higher education and higher income levels increased likelihood of seeking formal medical care. Trust in quality of care and shorter distances between patient points of origin and healthcare facilities increased the likelihood of seeking care from said facilities (Matsubara et al. 2019; Rasul et al. 2019). Having a chronic illness increased the probability of bypassing primary health centers (PHCs) among educated patients Li et al. (2021) in China. With regard to the few Indian studies that involved surveying patients physically present at healthcare facilities for recording HSB of patients, Chadda et al. (2001), Kaur et al. (2013), and Kapoor et al. (2019) analyzed treatment seeking practices of patients with respect to specific health conditions such as mental illnesses (Chadda et al. 2001) or tuberculosis (Kaur et al. 2013). These studies surveyed patients visiting only specialized healthcare facilities such as a psychiatric institution (Chadda et al. 2001) or a tertiary referral center (Kapoor et al. 2019) and reported gender-based differences in care seeking among patients. Rao & Sheffel (2018) conducted cross-sectional surveys in a rural Indian district and observed that patients bypassed primary care facilities for multiple reasons, including infrastructural deficiencies and a lack of trust in the quality of care.

We now discuss the survey methodology employed in previous studies assessing patient HSB. Thompson et al. (2016), Jacobs et al. (2017), and Matsubara et al. (2019) utilized both primary and secondary methods of data collection to record responses of participants. Lafort et al. (2016), and Habtu et al. (2018) conducted community-based cross-sectional surveys and longitudinal surveys to assess the HSB of their target population. Different sampling techniques such as stratified random sampling (Matsubara et al. 2019), systematic random sampling (Habtu et al. 2018), and response driven sampling (Lafort et al. 2016) were used for selecting study respondents. Both descriptive and inferential statistical tools were utilized to estimate the influence of socioeconomic and demographic variables on HSB of respondents. Thompson et al. (2016), Jacobs et al. (2017), and Rasul et al. (2019) used inferential analysis methods such as multiple logistic regression, multivariate tobit regression, probit modelling, multinomial and mixed conditional logit to estimate the influence of socioeconomic and demographic variables on respondent HSB. Low et al. (2016) qualitatively explored influence of social networks such as family members, friends, and healthcare providers on HSB of patients. We provide a detailed summary of the literature in Table 1a with the last row providing details about our current work.

**Table 1a:** Summary of previous literature.

| Study | Variables | Pilot survey | Standard sample size | Analysis | Survey type | Survey mode | Disease | Attributes | Place |
| --- | --- | --- | --- | --- | --- | --- | --- | --- | --- |
| Nemet & Bailey (2000) | A | Y | 390, N, PEN, SEN, RRN, ERN | $\chi^2$, MLR | CS | Mail | GH | Elderly | Vermont |
| Chadda et al. (2001) | G, A, EL, MS, E, FT | N | 78, N, PEN, SEN, RRN, ERN | $\chi^2$, FT | CS | HCF | PD | NA | Delhi, India |
| Gulati et al. (2009) | A, NC, EL, R, LT | N | 34034, N, PEN, SEN, RRN, ERN | BLR, MLR | CS | SD | RM | Women of 15 – 49 years | All Indian states |
| Yimer et al. (2009) | G, A, O, EL, MS | N | 14598, Y, PEN, SEY, RRN, ERN | BLR, MLR | CS | HS | Tuberculosis | Gender | Ethiopia |
| Narang, 2010 | G, EL, IL, A | N | 500, N, PEN, SEN, RRN, ERN | LR | CS | HCF | GH | NA | Lucknow, India |
| Nimbarte et al. (2011) | A, G, E, RT, IL | Y | 189, N, PEN, SEN, RRN, ERN | DA | CS | HCF | Tuberculosis | > 15 years | Maharashtra, India |
| Kaur et al. (2013) | A, AR, MS, EL | N | 109, N, PEN, SEN, RRN, ERN | $\chi^2$, $t$ | CS | HCF | Tuberculosis | Gender | Chandigarh, India |
| Kanungo et al. (2015) | A, G, R, C, EL, O, HO | N | 43999, N, PEN, SEN, RRN, ERN | BLR, MLR | CS | HS | GH | LI | West Bengal, India |
| Keesara et al. (2015) | A, EL, P | N | 61, N, PEN, SEN, RRN, ERN | QA | CS | HS | Post-partum | Women | Kenya |
| Lafort et al. (2016) | A, EL, RR | N | 1687, N, PEN, SEN, RRN, ERN | LR | CS | HS | HIV | FSW | 4 countries |
| Low et al. (2016) | A, G, EL, DD | N | 39, N, PEN, SEN, RRN, ERN | QA | CS | HCF | Diabetes mellitus | > 30 years | Malaysia |
| Tang et al. (2016) | A, G, EL, E, IL | N | 507, N, PEN, SEN, RRN, ERN | MLM, LCM | CS | HS | GH | Urban | China |
| Thompson et al. (2016) | A, G, IP, HM, TP | N | 7260, N, PEN, SEN, RRN, ERN | LR | CS | SD | Mental health | Gender | Canada |
| Awoke et al. (2017) | A, G, MS, EL, R, WQ, HI | N | 2517, N, PEN, SEN, RRN, ERN | MLR, BLR, $\chi^2$ | CS | SD | GH | Older adults | Ghana |
| Shahabuddin et al. (2017) | A, EL | N | 35, N, PEN, SEN, RRN, ERN | QA | CS | HS | Maternal HSB | MA | Bangladesh |
| Tenenbaum et al. (2017) | G | N | 3368, Y, PEN, SEY, RRN, ERN | $\chi^2$, MW, LR | CS | HCF | WT | Gender | Sweden |
| Habtu et al. (2018) | MS, E, R, A, EL, O, IL | N | 595, Y, PEN, SEY, RRY, ERN | LR | CS | HS | CC | CW | Ethiopia |
| Liu et al. (2018) | G, A, EL, IL, FS | N | 57, N, PEN, SEN, RRN, ERN | QA | CS | HS | Diabetes | CI | China |
| Minhas et al. (2018) | A | N | 2400, Y, PEN, SEN, RR, ERN | QA | CS | HS | GH | PUV | HP, India |
| Musinguzi et al. (2018) | G, RT, EL, MS, A, BP | N | 48, N, PEN, SEN, RRN, ERN | QA | CS | HS | Hypertension | NA | Uganda |
| Rao & Sheffel (2018) | G, A, WQ, LL | N | 11981, N, PEN, SEN, RRN, ERN | LR | CS | HCF, HS | GH | Sick patients in past 30 days | Chhattisgarh, India |
| Welay et al. (2018) | G, E, R, MS, OC, EL | N | 423, Y, PEN, SEN, RRY, ERN | MLR | CS | HS | GH | NA | Ethiopia |
| Kapoor et al. (2019) | G, A, SOR | N | 237708, N, PEN, SEN, RRN, ERN | LR | CS | HCF | GH | Gender | Delhi, India |
| Lim et al. (2019) | S, A, EH, EL, HI, ES | N | 3979, N, PEN, SEN, RRN, ERN | MLR | CS | HCF | GH | PC | Malaysia |
| Matsubara et al. (2019) | A, O, HAI, EL | N | 315, N, PEN, SEN, RRN, ERN | LR | CS | HS | Delivery | Poor | Vietnam |
| Ngangbam & Roy, 2019 | G, A, MS, EL, R, HS | N | 40564, N, PEN, SEN, RRN, ERN | PM, MLR, MCL | CS | SD | GH | NA | Northeast India |
| Rasul et al. (2019) | A, G, MS, O, HH | N | 6958, N, PEN, SEN, RRN, ERN | MLR | CS | HS | NCD | NA | Bangladesh |
| You et al. (2019) | A, EL, MS, R, HI | N | 14316, PEN, SEN, RRN, ERN | FLR | CS | HS | KHPS | Gender | Korea |
| Deolia et al. (2020) | G, A | Y | 700, N, PEN, SEN, RRN, ERN | $\chi^2$ | CS | HS | Oral health | Rural | Maharashtra, India |
| Marsh et al. (2020) | A, EL, R, FS, LT | N | 750, N, PEY, SEN, RRY, ERN | MLR | LS | HS | CI | Women | Pune, India |
| Ndarukwa et al. (2020) | G, A, EL, E, MS, R, AR | N | 400, Y, PEN, SEN, RRN, ERN | BA, MLR | CS | HCF | Asthma | Adult | Zimbabwe |
| Rana et al. (2020) | G | N | 13244, N, PEN, SEN, RRN, ERN | $\chi^2$, MLR, $t$ | Review | No | Lung cancer | Gender | Australia |
| Campbell et al. (2021) | G | N | 566829, N, PEN, SEN, RRN, ERN | MLLR | CS | SD | PD | Gender | 73 countries |
| Srivastava et al. (2021) | A, G, EL, MS, IL | N | 31464, N, PEN, SEN, RRN, ERN | $\chi^2$, HPSM, LR | CS | SD | PD | Older | All Indian states |
| Thomas et al. (2021) | A, G, O, EL, BMI | N | 74532, N, PEN, SEY, RRY, ERN | $\chi^2$, MLR | CS | HS | TB | Gender | 17 Indian states |
| **Current work** | **G, A, MS, EL, O, IL, NC** | **Y** | **450, Y, PEY, SEY, RRY, ERY** | **BLR, MLR** | **CS** | **HCF** | **GH** | **Urban** | **New Delhi, India** |

**Table 1b:** Acronyms.

| Variables |  | Sample size | Analysis |
| --- | --- | --- | --- |
| A: Age | HO: House ownership | Y: Yes | DA: Descriptive analysis |
| AR: Area of residence | C: Caste | N: No | $\chi^2$: Chi squared test |
| MS: Marital status | BMI: Body mass index | PEY: Population size effect considered | $t$: t test |
| EL: Education level | SOR: State of Residence | PEN: Population size effect not considered | BLR: Binomial logistic regression |
| R: Religion | P: Parity | SEY: Sampling effect considered | MLR: Multinomial logistic regression |
| O: Occupation | WQ: Wealth quantile | SEN: Sampling effect not considered | MW: Mann Whitney test |
| IL: Income level | NC: Number of children | RRY: Response rate considered | MLM: Mixed logit model |
| LT: Locality type | HS: Housing status | RRN: Response rate not considered | LCM: Latent class model |
| RT: Residence type | MHSB: Maternal HSB | ERY: Eligibility rate considered | CLM: Conditional logit model |
| IP: Illness prevention | MA: Married adolescents | ERN: Eligibility rate not considered | BA: Bivariate analysis |
| HM: Health maintenance | LI: Low Income |  | HPSM: Heck probit selection model |
| TP: Trust in physician | Pilot survey | Survey mode | LR: Logistic regression |
| FS: Family structures | Y: Yes | TS: Telephonic survey | FT: Fisher test |
| E: Ethnicity | N: No | HCF: Healthcare facility-based survey | MLLR: Multi level linear regression |
| DD: Disease duration | Survey type | HS: Household based survey | FLR: Fractional logit regression |
| RT: Residence type | LS: Longitudinal survey | SD: Secondary data | PM: Probit model |
| BP: Blood pressure level | CS: Cross sectional survey | Disease | MCL: Mixed conditional logit |
| FS: Family size | Place | G&RI: Gastrointestinal and respiratory illness | NCD: Non-communicable diseases |
| E: Employment | HP: Himachal Pradesh | GH: General health | WT: Whiplash trauma |
| FT: Family type | CW: Childbearing women | PD: Psychiatric disorder | CI: Childhood Illness |
| CC: Cervical cancer | FSW: Female sex workers | RM: Reproductive Morbidity | PUV: Parents of under five |
| LL: Literacy level | PC: Primary care | CI: Chronic illness |  |

With respect to previously published work on HSB, we collect patient HSB data from multiple primary and secondary public healthcare facilities based in a metropolitan city to determine significant factors influencing patients to visit public healthcare facilities. Collecting HSB information via in-person visits to healthcare facilities provides authentic and reliable information about participants actually accessing the system. We contribute to the literature by analyzing overarching patient HSB without limiting the study to a specific disease, and further focus on quantifying the association of multiple socioeconomic and demographic attributes of patients such as gender, age, education level, annual income, marital status, employment status, and number of children with their HSB. In addition to collecting socioeconomic and demographic information of participants, we also recorded their travel times to the facility as well as their modes of transportation, which were not recorded in previous studies in urban settings.

In our knowledge, none of the previous studies estimated the study sample size based on a pilot survey, and neither did they consider incorporating the effects of response rate, inclusion rate, and study design in determining the final sample size for the survey. In particular, in the Indian context, the majority of the existing literature also did not provide a detailed quantitative analysis of participant responses. Thus our study provides a quantitatively rigorous analysis of patient HSB and their socioeconomic and demographic determinants in the context of urban public primary and secondary healthcare delivery.

## 2. Methodology

In this section, we describe the survey design, conduct and analysis approach. We first describe the primary and secondary public healthcare facilities where the survey was conducted. We then explain the survey design and the survey sample size estimation procedure. We then briefly discuss the modelling technique used to determine the significant factors associated with patient HSB.

### 2.1 Healthcare facilities description

We conducted the cross-sectional survey at twenty-two primary and secondary public healthcare facilities from December 2019 to April 2022 in the South-west Delhi district in India. South-west Delhi is one of the eleven administrative districts of National Capital Territory of Delhi in India with seventy-seven villages in the district (Delhi, 2018). We present South-west Delhi district profile in Table 2. We chose South-west Delhi as the study area owing to two reasons: (a) availability of significantly larger number of primary and secondary public healthcare facilities in comparison to other districts, and (b) proximity to the authors’ institute. We had to suspend the survey twice due to the COVID-19 pandemic and gathered participant responses within a total duration of six months.

**Table 2:**
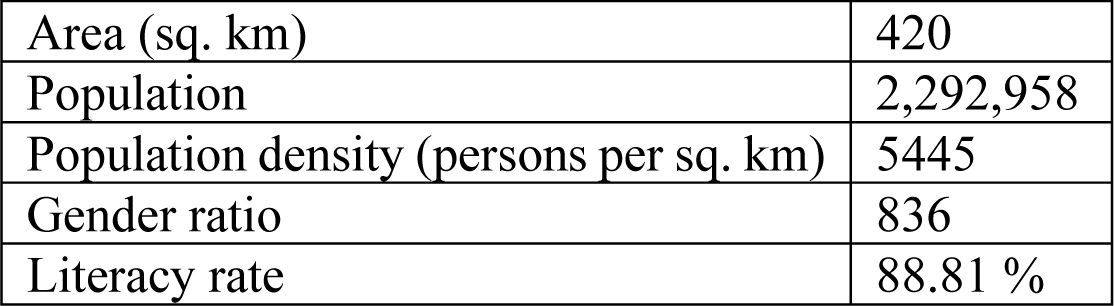
South-west Delhi district profile.

|  |  |
| --- | --- |
| Area (sq. km) | 420 |
| Population | 2,292,958 |
| Population density (persons per sq. km) | 5445 |
| Gender ratio | 836 |
| Literacy rate | 88.81 % |

We made in-person visits to three types of healthcare facilities - dispensaries, primary urban health centers (PUHCs), and polyclinics - categorized under primary and secondary level of care under the Delhi public healthcare delivery system (Department of Health & Family Welfare, 2020). We collected key operational details such as services offered at different facilities, average patient load per day at different departments, availability of different types of healthcare providers, and other information based on discussion with doctors and data collected from patient records maintained at these facilities.

The Government of NCT of Delhi has a three-tier healthcare delivery system including dispensaries, PUHCs, polyclinics, secondary hospitals, and tertiary care hospitals to provide healthcare services to its target population (Department of Health & Family Welfare, 2020). Primary healthcare services are available to patients through dispensaries and PUHCs. Polyclinics have been set up for providing specialized services through specialists in medicine, paediatrics, ophthalmology, orthopaedic, gynaecology, ENT, and dermatology. Facilities offering a higher level of care than polyclinics provide a wide range of services across clinical specialties such as surgery, cardiology, nephrology, and urology.

Dispensaries provide treatments of simple ailments including fever, mild infections, first aid and management for wounds, and also provide basic diagnostic facilities. Along with outpatient consultations with general physicians, dispensaries also offer antenatal and prenatal care services (ANC and PNC) to patients, and immunization services to children below age of 10 years. In terms of availability of healthcare providers, there are one to two general physicians, one to two ANC nurses, one laboratory technician, one pharmacist, and a receptionist for registering patient information. There are twenty-three dispensaries in South-west Delhi and a single dispensary aims to cater to a population of 30,000 – 50,000 persons within a radius of 5 kilometers (kms). From our visits, we observed significant variation in the average number of patients visiting dispensaries each day. For instance, the minimum and maximum daily average outpatient loads across dispensaries were 60 and 200 with an average of 120 patients. This significant variation in patient load leads to inequitable utilization of medical resources, resulting in overcrowding at a few dispensaries in comparison to others.

Next, we visited PUHCs, which aim to serve approximately 50,000 persons. PUHCs are mandated to offer free outpatient department (OPD) services, ANC and PNC services and other services including family welfare, immunization, diagnostic, dental, geriatric care, gynaecology, and tuberculosis treatment. However, based on our visits, we observed patient flows and other operational services at PUHCs to be similar to those of dispensaries without provision of dental and geriatric care facilities. In terms of medical resources, there are one to two general physicians, one to two nurses for attending to ANC and PNC patients, one pharmacist, lab technician, and a receptionist. The average daily outpatient load at PUHCs varies between 120 – 140 patients.

We also administered surveys at polyclinics, which are meant to provide healthcare at the secondary level. Per government guidelines, polyclinics are specialist outpatient clinics where medicine, gynaecology, and paediatrics specialists are supposed to be available every day and orthopaedics, dermatology, ophthalmology, and ENT specialists are supposed to be available on selected days of the week. However, during our visits, we observed that gynaecology and paediatrics services are also provided only on a few days of the week. We observed that the general medicine OPD is operational all six days a week, paediatric care is provided on Mondays and Fridays, obstetrics and gynaecology on Tuesdays and Saturdays, surgery on Tuesdays, orthopaedics on Thursdays, ophthalmology services on Fridays, and ENT services on Saturdays. Similar to the functioning of dispensaries, polyclinics are also operational until 2 PM. We observed significantly higher average patient loads per day at polyclinics in comparison to dispensaries and PUHCs. We summarize the operational characteristics from two representative dispensaries, PUHCs, and polyclinics at South-west Delhi in Table 3.

**Table 3:** Operational configurations of primary and secondary public healthcare facilities. OPD = outpatient department; ENT: ear, nose and throat; ANC = antenatal care; PUHC = primary urban healthcare centers.

| Healthcare facility type |  | Average daily OPD load, general medicine | Specialist services: number of specialists (daily average patient load) |  |  |  |  |  |  | ANC services | Immunization services | Average number of general physicians, lab technician, receptionist, pharmacist, ANC nurse |
| --- | --- | --- | --- | --- | --- | --- | --- | --- | --- | --- | --- | --- |
|  |  |  | Gynaecology | Ophthalmology | Orthopaedics | Paediatrics | Dermatology | ENT | Surgery |  |  |  |
| Dispensaries (23) | 1 | 190 | NA |  |  |  |  |  |  | 40 – 50 | 45 – 55 | 2, 1, 1, 1, 2 |
|  | 2 | 85 | NA |  |  |  |  |  |  | 15 – 40 | 20 – 40 | 1, 1, 1, 1, 1 |
| PUHCs (7) | 1 | 130 | NA |  |  |  |  |  |  | 50 – 60 | 60 – 70 | 2,1,1,1, 1, 2 |
|  | 2 | 100 | 1 (60 – 70) | NA |  |  |  |  |  | 40 – 50 | 50 – 60 | 1,1,1,1,1, 1 |
| Polyclinics (4) | 1 | 210 | 1 (60 – 70) | 1 (60 – 80) | 1 (100 – 110) | 1 (70 – 80) | 1 (120 – 130) | 1 (80 – 85) | 1 (40 – 45) | 50 – 60 | 60 – 70 | 2,1,1,1, 1, 2 |
|  | 2 | 160 | 1 (20 – 30) | 1 (70 – 90) | 1 (120 – 150) | 1 (80 – 100) | 1 (75 – 80) | 1 (80 – 90) | 1 (30 – 40) | 50 – 60 | 60 – 70 | 1,1,1,1, 1, 2 |

Information collected from in-person visits and consultations with doctors at the facilities.

### 2.2 Survey questionnaire design and description

We designed survey questionnaires for patients in the English and Hindi languages. We divided the survey questionnaire into 2 sections with 15 questions enquiring about: (a) HSB (including reasons leading to healthcare seeking, types of healthcare services utilized, and average time to reach healthcare facility, among others - 8 questions), and (b) demographic details (7 questions). Demographic details collected included patient gender, age, marital status, annual income, number of children, employment status, and education level. We present the first part of the survey questionnaire in Figure 1.

**Figure 1:**
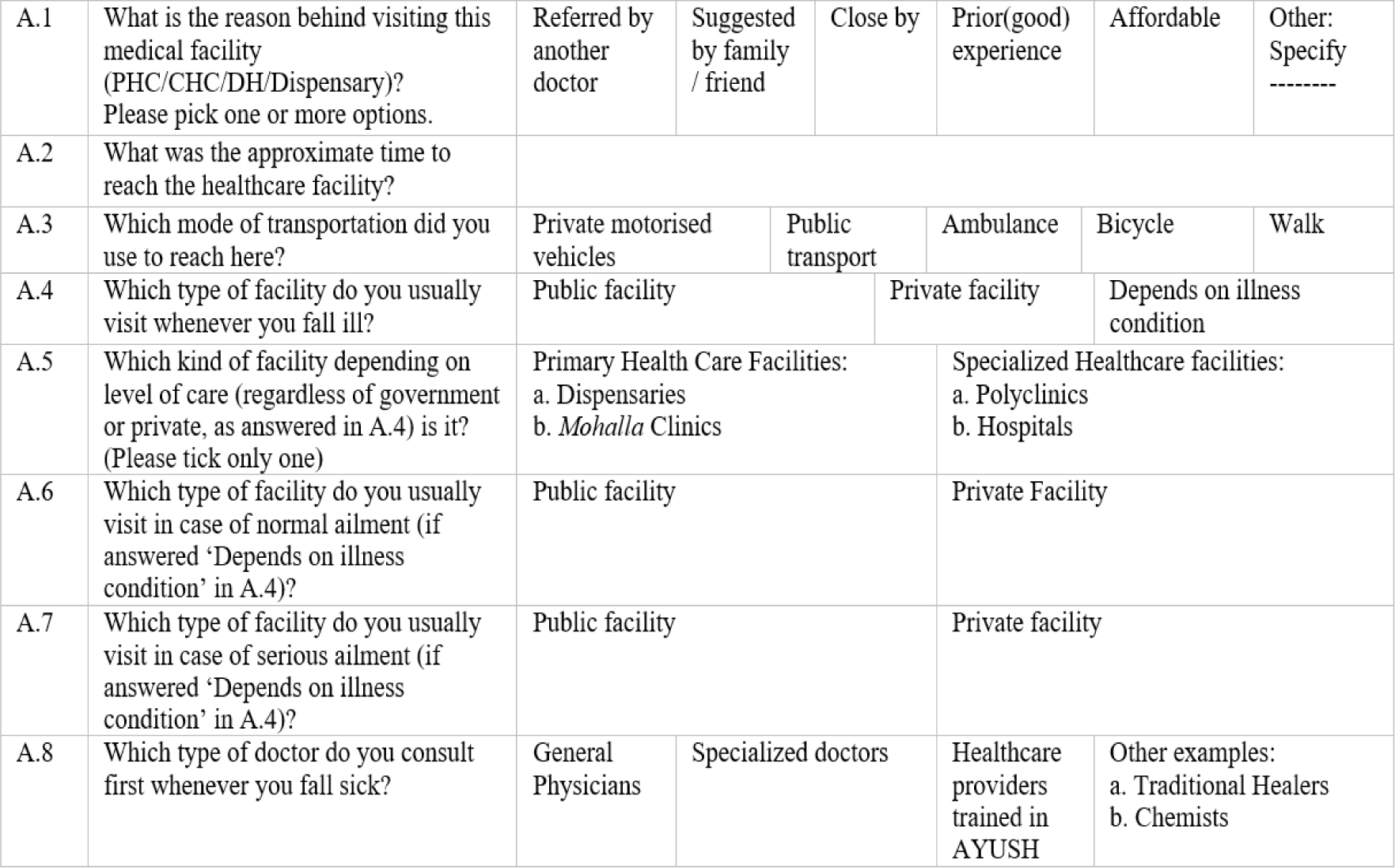
Survey questionnaire for recording HSB of participants. Note: AYUSH (Ayurveda, Yoga and Naturopathy, Unani, Siddha, Homeopathy).

We briefly discuss questions presented in Figure 1 meant to examine HSB of patients. First, we explored motivating factors for seeking care from primary and secondary public healthcare facilities in a large metropolitan city such as New Delhi, wherein a large number of private alternatives are available. We allowed participants to choose more than one option in their response to this question (question A.1). Next, we recorded the average time it took patients to reach the healthcare facility using different modes of transportation (questions A.2 and A.3) in order to understand how accessible these facilities are to the general population.

First, we explored motivating factors for seeking care from primary and secondary public healthcare facilities – dispensary, PUHC, or polyclinics in a large metropolitan city like Delhi with availability of significantly large number of private alternatives. We allowed participants to choose more than one option in their response to this question (question A.1). Next, we recorded the time it took patients to reach the healthcare facility using different modes of transportation (questions A.2 and A.3) in order to understand how accessible these facilities are to the general population.

Subsequently, via question A.4, we attempted to determine the type of healthcare facility patients – public or private - preferred to visit first upon falling ill. We note here that while public healthcare facilities offer free medical services for every resident per the guidelines of the Government of Delhi (DGHS, 2017), private healthcare facilities operate autonomously and provide medical care on a chargeable basis with significant variance in both quality of care provided and the fees charged (Ram, 2021). For patients who indicated that their preference for visiting a particular type of healthcare facility was dependent on their illness condition, we recorded their responses separately via questions A.6 and A.7 depending upon their perception of the severity of their illness.

We next examined, via question A.5, what level of healthcare facility – primary care or more specialized facility, regardless of whether they are public or private facilities - patients preferred to visit first upon falling ill. Under primary care facilities, we included ‘*mohalla’* (neighbourhood in Hindi) clinics or dispensaries, and we included polyclinics and hospitals under specialized facilities. This question was meant to quantitatively determine the likelihood of patients in an urban metropolitan city such as New Delhi to bypass lower-level facilities for seeking care directly at specialized facilities. Finally, via question A.8, we attempted to determine the preference of patients for a specific type of doctor to visit upon first falling ill (general physicians, specialized doctors, doctors formally trained in traditional forms of medicine, and others including informal traditional healers and chemists), regardless of which type of facility the provider is situated in (public/private, primary/higher level of care).

### 2.3 Survey data collection

We conducted in-person face-to-face interviews using a close-ended survey questionnaire from respondents present at healthcare facilities. We asked respondents to choose from the available options in the questionnaire by ticking or encircling their desired category. Participation was voluntary, and respondents indicated their willingness to participate in a consent form attached to each questionnaire. We observed an overall response rate of ninety percent. We read out questions to patients during the interview and ensured that participation of respondents to survey did not affect regular healthcare delivery operations at healthcare facilities. Initially we conducted a pilot survey among forty respondents and based on participant responses, we modified and prepared the final version of the questionnaire. We did not collect any personally identifiable information from patients.

### 2.4 Survey administration methodology

We considered patients eligible to participate in the survey if they were: (a) aged greater than 18 years, (b) not severely ill and therefore not in a position to respond, and (c) idle and waiting for their consultation in the queue. We assigned numerical codes to each response category included in the survey questionnaire and compiled participant responses in a single Microsoft Excel sheet. In case of discrepancies or missing responses, we discarded the entire response of the particular respondent.

### 2.5 Ethics statement

The Institutional Review Board at the Indian Institute of Technology New Delhi approved the study protocol with approval number – IITD-IEC-ID-P064 on November 25, 2019. We asked each potential respondent to sign on the participant informed consent form (PICF) prior to the data collection process. We wrote names of the illiterate patients in the PICF on their behalf after explaining the relevant study details.

### 2.6 Sample size estimation

We estimated the sample size for the survey based on pilot survey responses from forty participants. For each question *N*_1_, *N*_2_, *N*_3_,….., *N*_*s*_ included in the questionnaire, we first estimated its corresponding sample size depending upon whether the response variable for the question was a discrete or continuous variable (Aday & Cornelius, 2006).

For questions with continuous response variables, the sample size *S*_*C*_ was estimated using equation 1 below.

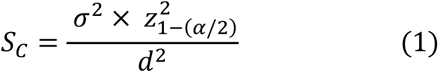

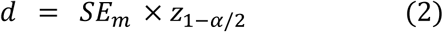

Here, *σ* is the standard deviation estimated for participant responses during the pilot survey for each question with a continuous response variable, *z*_1-(*α*/2)_ is the standard normal random variable quantile associated with a level of significance *α, d* is the margin of error and *SE*_*m*_ is the standard error of the mean.

For questions with discrete response variables, the sample size *S*_*d*_ was estimated using equation 3 below.

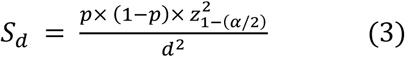

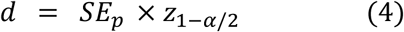

Here, *p* is the proportion estimated for each category of the discrete response variable during the pilot survey, and *SE*_*p*_ is the standard error of the proportion estimate *p*.

After calculating *S*_*C*_ and *S*_*d*_, we estimated additional factors such as the population size effect, sampling effect, response rate, and eligibility rate using expressions provided below and incorporated all these factors in estimating the final sample size for the survey.

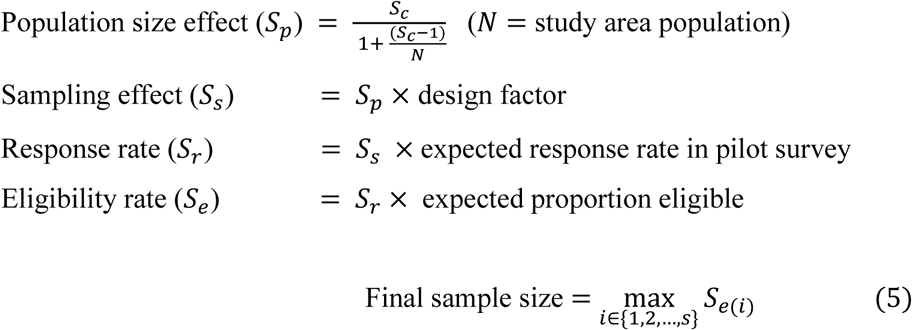

We repeated the above calculations for each question included in the survey questionnaire and then estimated final sample size using equation 5. The final sample size for the patient survey was found to be four hundred and forty-nine.

### 2.7 HSB modelling

We analyzed the relationship between patient HSB and their socioeconomic and demographic attributes using two logistic regression techniques: (a) binomial and (b) multinomial, depending upon number of categories of response variables. We provide the list of dependent variables for analyzing HSB of patients along with associated reference categories in Figure 2. We did not perform logistic regression for the data collected for question A.1 provided in Figure 1 because the majority of patients chose more than one option in their response.

**Figure 2:**
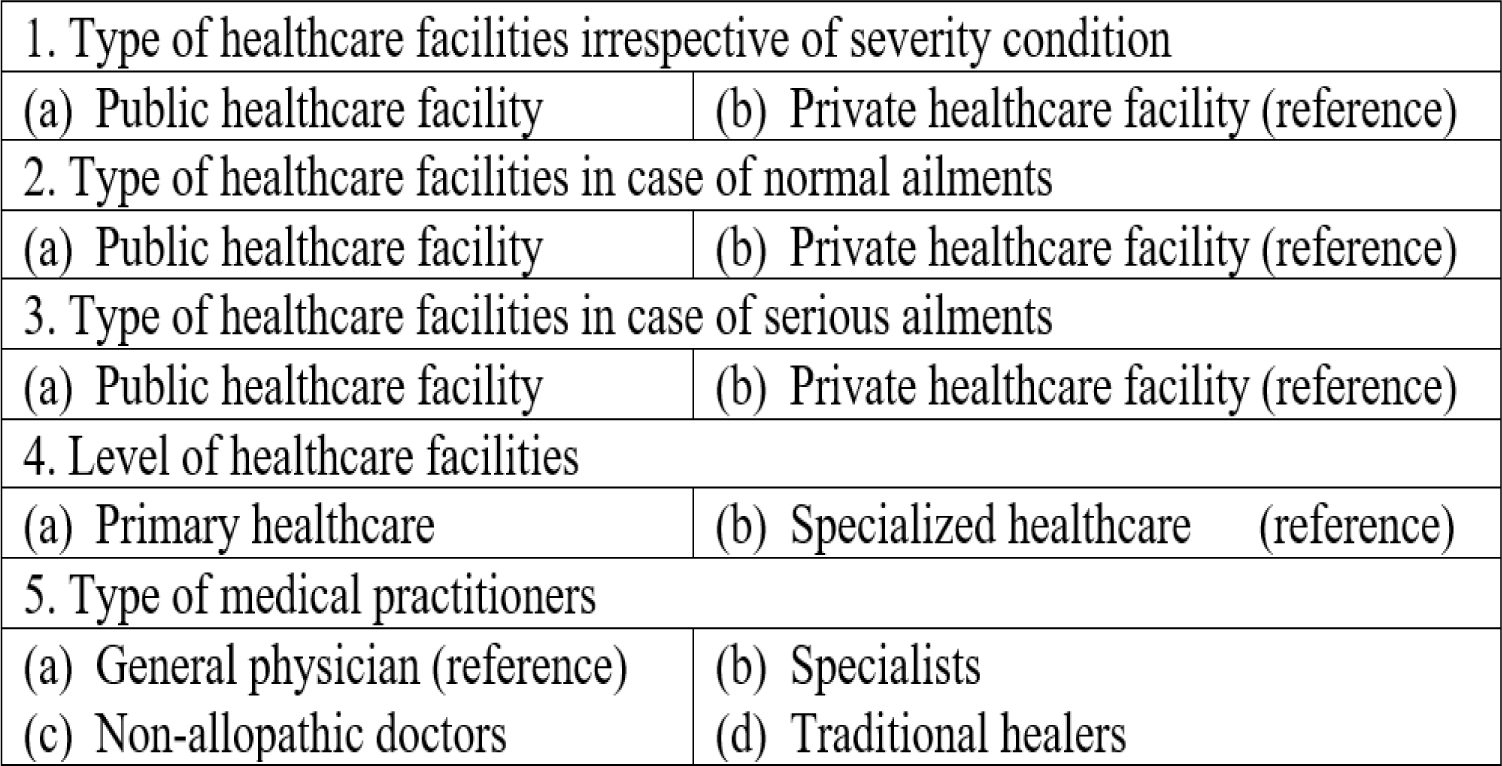
List of dependent variables for logistic regression with reference categories.

We developed five logistic regression models: four binomial (1-4 in Figure 2 above) and one multinomial (5 in Figure 2) for analyzing HSB of patients. Independent variables were patient attributes including gender, age, marital status, education level, annual income level, occupational status, and number of children. The logistic regression analyses were performed on the R statistical computing platform (Venables & Smith, 2022).

## 3. Results

We now quantitatively describe HSB of patients in urban context based on survey responses. We begin by describing the profile of the surveyed population and their HSB via descriptive statistical analyses.

### 3.1 Descriptive analysis

We provide a summary of the surveyed population in Table 4. In our surveyed sample, the majority of patients were female (70.67%) and married (77.56%). In terms of education, approximately 18.88% of patients did not receive any form of formal education and among patients with formal education, almost a third were undergraduates. The highest proportion of patients visiting public healthcare facilities were homemakers (38.44%) followed by professionally employed persons (36.67%). A significant proportion of patients (24.22%) preferred not to reveal their income level.

**Table 4:**
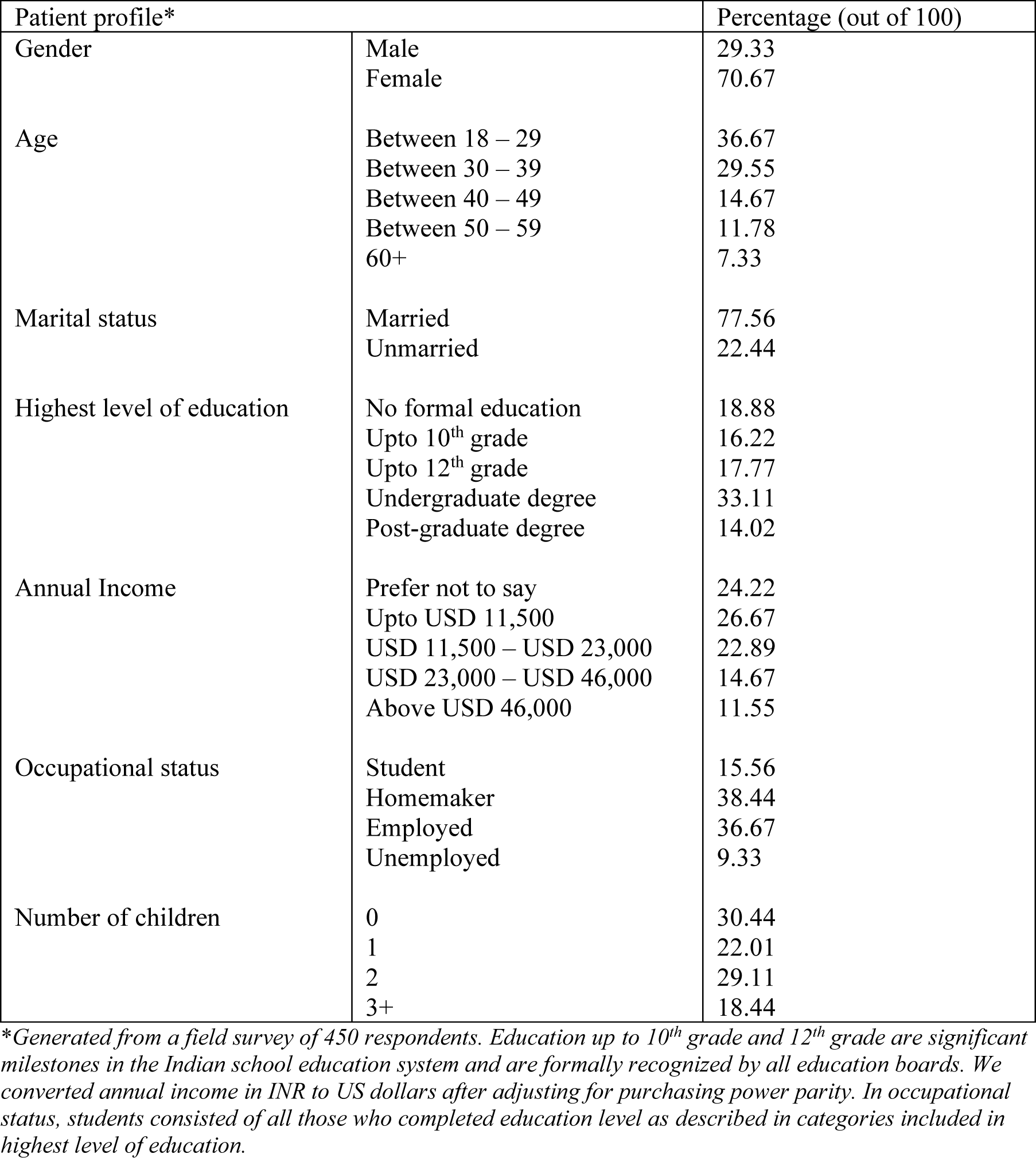
Participant profile characteristics at South-west Delhi public healthcare facilities.

With regard to reasons why patients visited the primary or secondary public healthcare facilities where we conducted the survey (question A.1 in Figure 1), good prior experiences in terms of provider attitudes and trust in the quality of medical services motivated a significant proportion of patients (62.22%) to visit these healthcare facilities. Other reasons including cleanliness, lesser wait times at healthcare facilities, and provision of free laboratory services prompted patients to return to the same facility. We also observed proximity to healthcare facilities from the patient’s residence increased the likelihood of seeking care from a given health facility (Figure 3). Similar observations were confirmed by Ndarukwa et al. (2020) studying HSB of adult patients with asthma at a public hospital in Zimbabwe. Second, we investigated the means of transportation used by patients to arrive at healthcare facilities. The highest proportion of patients (54.22%) walked to the health facility, which may indicate reasonably quick accessibility to primary and secondary care within the existing healthcare delivery system in the region studied. We observed from Figure 4a that a small proportion of patients (approximately 1.55%) were carried on ambulances to healthcare facilities. These ambulance services were arranged by the patients themselves and were not provided by these facilities as it is not part of their service mandate.

**Figure 3:**
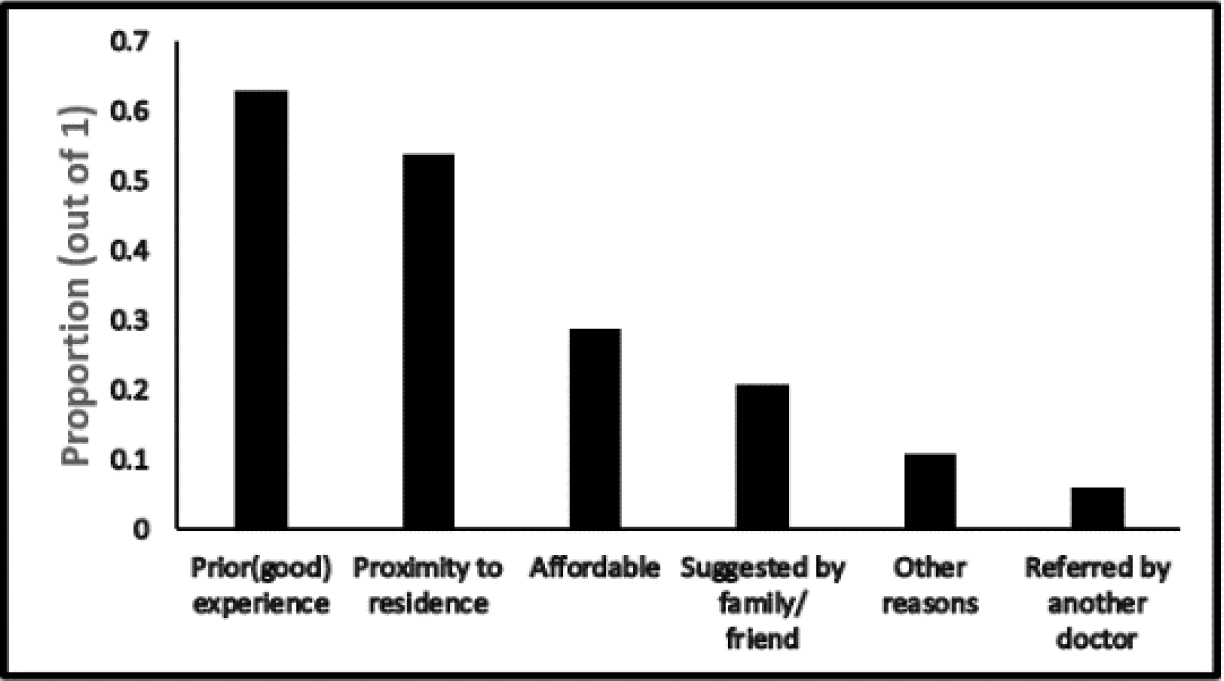
Reasons for visiting public healthcare facilities.

**Figure 4a:**
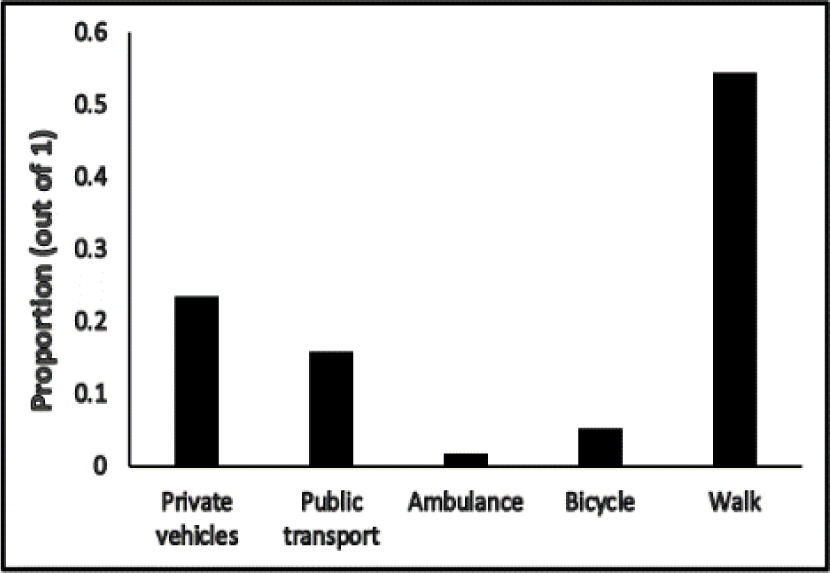
Different modes of transportations.

Via the survey, we also estimated the average travel time it took patients to reach the primary and secondary healthcare facilities under consideration, and further categorized it on the basis of the various modes of transportation patients used. The reported travel time to reach health facilty was lowest for patients visiting healthcare facilities in private motorized vehicles (mean = 8.76 minutes, standard deviation [SD] = 4.75 minutes), followed by patients with bicycles (mean = 12.09 minutes, SD = 5.24 minutes). We summarize the average travel time statistics in Figure 4b, where we observe patients visting healthcare facilities via public transport took the longest time (mean = 15.77 minutes, SD = 10.17 minutes) to reach the health facility.

**Figure 4b:**
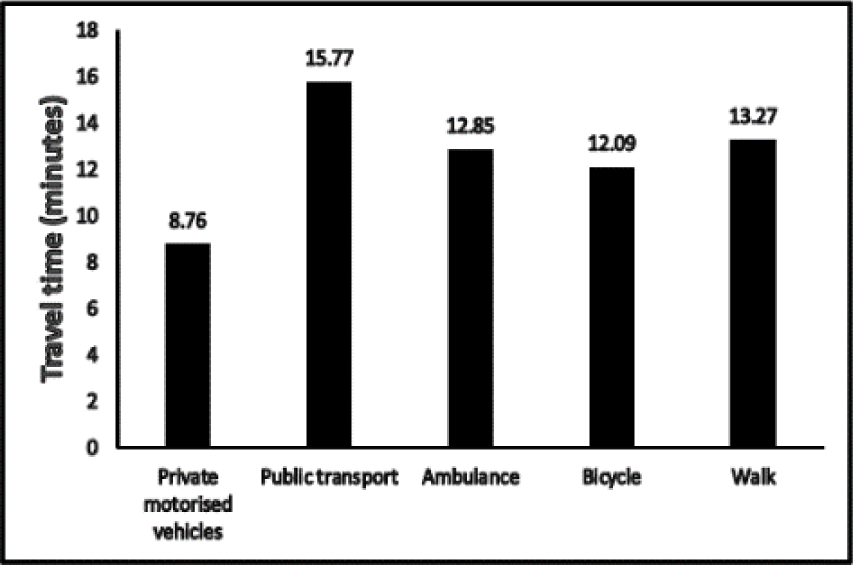
Average travel time to reach healthcare facilities.

Next, we determined the preferences of patients for public and private healthcare facilities. A significant proportion of patients (49.11%) favored visiting public healthcare facilities for any health-related issue regardless of their perceived severity of the disease condition. This finding is consistent with previous literature studying preferences of patients for seeking care from public healthcare providers in developing countries (E. Jia et al., 2020; Tang et al., 2016). Approximately 30.22% chose the healthcare facility on the basis of the perceived severity of their illness. For illnesses of perceived mild severity, a significantly larger proportion of patients (77.20%) frequently visited public healthcare facilities. For illnesses of perceived high severity and for chronic illness, 80.14% of the patients surveyed sought care from private healthcare facilities. Upon further enquiry, we found that shorter wait times before admission, especially at specialized levels of care, was the primary factor that influenced patients in their choice of private facilities, even though medical services were provided at significantly higher costs in these facilities. We report the inclination of patients towards different types of healthcare facilities in cases of illnesses of perceived mild and high severity in Figure 5a.

**Figure 5a:**
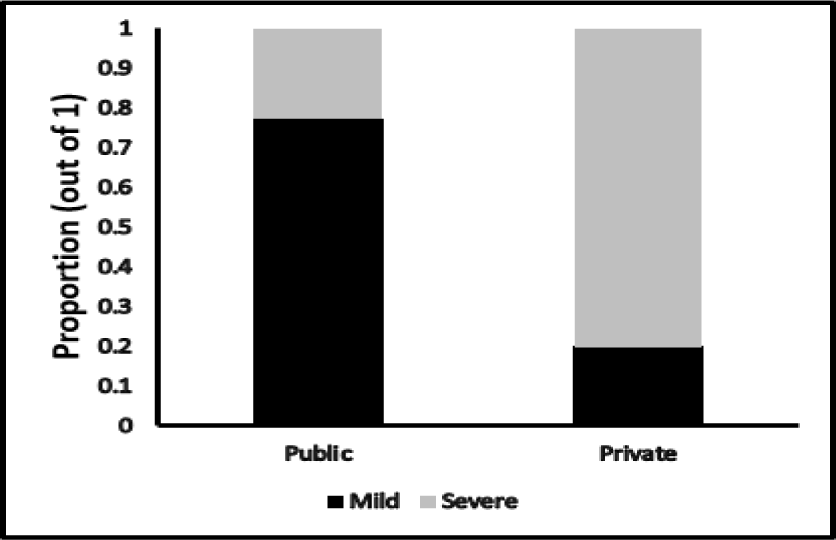
Preferences for different types of healthcare facilities for a first visit.

**Figure 5b:**
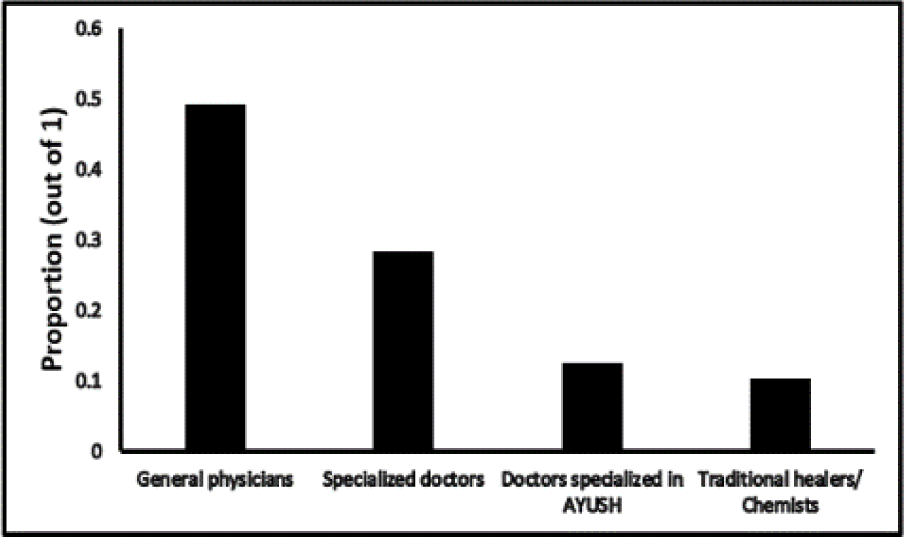
Patient preferences for different types of providers for the first visit. AYUSH: Ayurveda, Yoga and Naturopathy, Unani, Siddha, and Homeopathy.

Next, we recorded patient preferences for visiting: (a) primary level healthcare facilities, and (b) higher level, often specialized healthcare facilities. Regardless of the perceived severity of their illness, a substantial proportion of patients (40.44%) preferred to make a first visit to specialized healthcare facilities including clinics and hospitals. This behaviour of not following the hierarchy of the healthcare delivery system, having (sometimes undue) preferences for specialist services, and the absence of effective referral and gatekeeping mechanisms has been shown to cause delayed care and longer overall length of stays for patients requiring emergency services at specialized level of care (Kelen et al., 2021). We also examined patient preferences for consulting different types of healthcare providers. Most patients (49.11%) chose to consult general physicians followed by specialized doctors (28.22%) upon first feeling unwell (Figure 5b).

Interestingly, 10.22% of patients also sought care from traditional healers and among these respondents, majority (2.5 times higher) were female patients without any formal education (41.73%). Such healthcare practices indicate limited health literacy and lower trust in formal care among patients. A small proportion of patients reported that they consulted traditional healers after not finding relief from treatments suggested by formally trained healthcare providers.

### 3.2 HSB: Inferential analysis

In this section, we present inferential analyses of our survey results, which involves using binomial logistic regression (BLR) and multinomial logistic regression (MLR) models to determine relationships between socioeconomic and demographic variables and HSB. All analyses were carried out with a level of significance of 0.05. Prior to conducting the logistic regression analyses, we estimated the correlation matrix for the independent variables to assess the presence of multicollinearity. Based on the correlation matrix, we observed that marital status was highly correlated with age, employment status, and number of children, with correlation estimates of 0.48, 0.58, and 0.77, respectively. While the reported correlation estimates are significant, they are below the generally agreed rule of thumb criterion used in Midi et al. (2010) and Senaviratna & A. Cooray, (2019) that outline that multicollinearity may be considered to be significant if the correlation coefficient for two variables exceeds 0.8. The authors proposed addressing such multicollinearities by omitting one of the correlated variables from the regression model and then checking how the Akaike’s information criteria (AIC) scores change. Going by this approach, we observed relatively small differences in AIC scores and the estimates of the statistically significant parameters. We report results from the most parsimonious models along with the percentage change in AIC scores in Tables 5 and 6 for the BLR and MLR analyses, respectively.

**Table 5:**
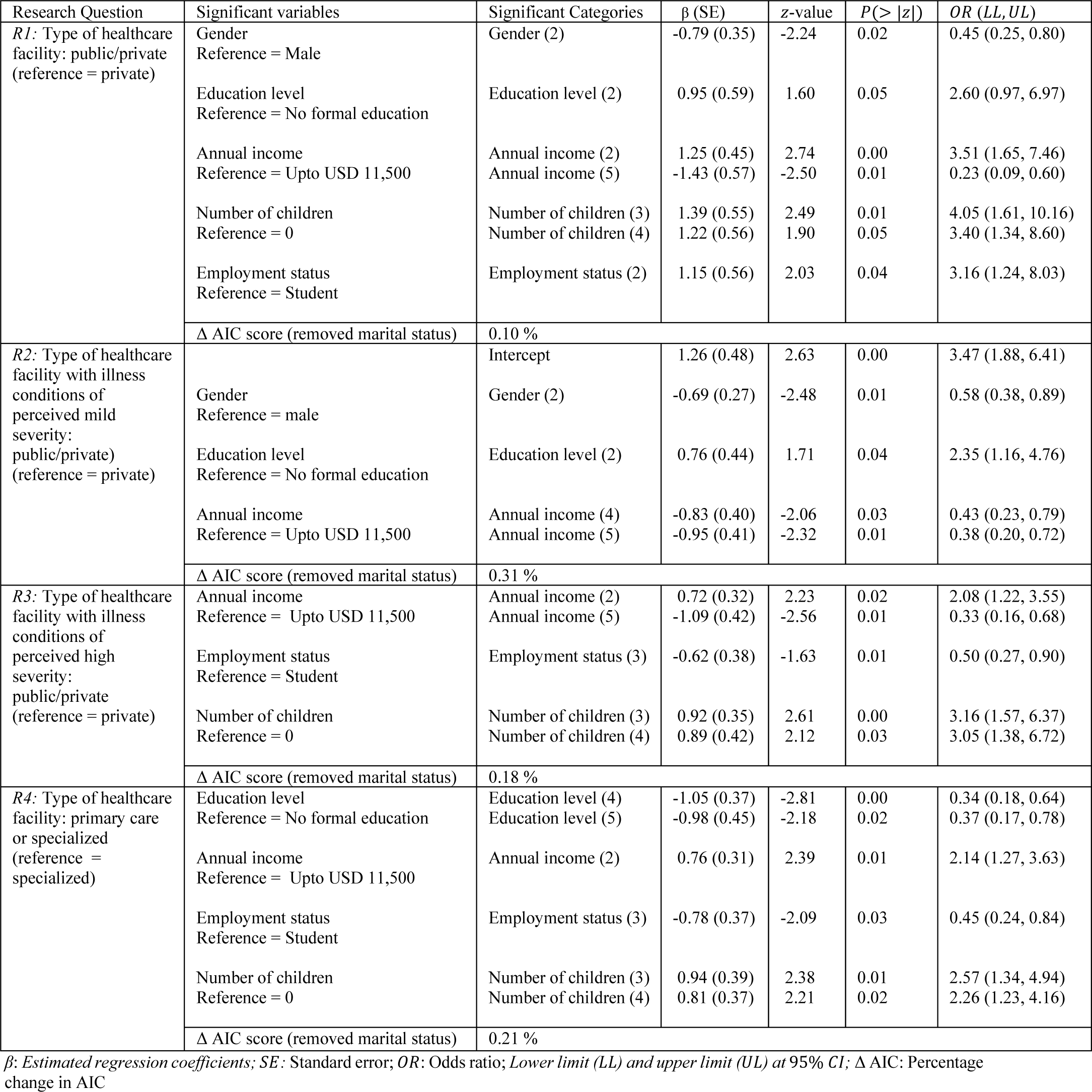
Results from BLR modelling of HSB as a function of socioeconomic and demographic variables.

#### 3.2.1 BLR based HSB model

Before conducting the BLR analyses, we formulated the research question and corresponding research hypotheses associated with each model. We present each such formulation below.

##### Research question (R1)

What socioeconomic and demographic factors are associated with a patient visiting a public or a private healthcare facility?

The null and alternate hypotheses associated with the above research question are as follows.

H_0_: Socioeconomic and demographic variables do not influence a patient’s choice of visiting a public or a private healthcare facility.

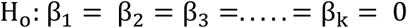

H_1_: Socioeconomic and demographic variables influence a patient’s choice of visiting a public or a private healthcare facility.

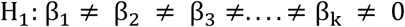

To answer *R1*, we first selected the reference categories for the predictor and response variables. The response variable had two categories: (a) public healthcare facility (labelled 1), and (b) private healthcare facility (labelled 0). We chose ‘private healthcare facility’ as the reference category, with the dependent variable being whether a person visits a public or a private healthcare facility given the set of attributes in Table 6. We treated all predictor variables as discrete, including patient age and annual income. We observed that five out of seven variables, i.e., (a) gender, (b) education level, (c) employment status, (d) annual income, and (e) number of children were significantly influencing patient choices. We present parameter estimates of significant independent variables in Table 5.

Thus the BLR model with significant socioeconomic and demographic predictor variables is given in equation (6) below.

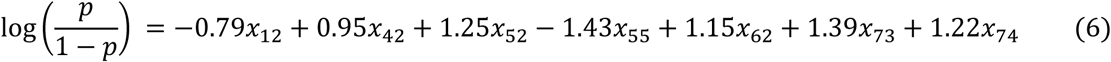

*p* = probability of visiting a public healthcare facility

*x*_12_ = 1, if the respondent is female

*x*_42_ = 1, if the respondent has completed 10^th^ grade

*x*_52_ = 1, if the respondent has annual income between USD 11,500 – USD 23,000

*x*_55_ = 1, if the respondent has annual income above USD 46,000

*x*_62_ = 1, if the respondent is homemaker

*x*_73_ = 1, if the respondent has 2 children

*x*_74_ = 1, if the respondent has 3 or more than 3 children

Based on the parameter estimates provided in Table 5 and equation 6, we observed that patients with higher annual income were more likely to visit private healthcare facilities in comparison to public healthcare facilities irrespective of the perceived severity of their illness condition. Relatively, patients with annual income above USD 46,000 were 4.34 times more likely to visit private healthcare facilities in comparison to patients with annual income upto USD 11,500. Further, we also observed that patients with higher education levels were less likely to visit a public healthcare facility (0.38 times less likely) in comparison to patients with no formal education. Similar observations were reported in other lower and middle income countries (Awoke et al. 2017; Basu et al. 2012; Wellay et al. 2018). Further, this was corroborated by patient comments during the survey, where they indicated that they also preferred to visit a private healthcare facility because of shorter waiting times before receiving care. We also observed that public healthcare facilities were 3.40 times more favored by patients having three or more than three children in comparison to patients with no children.

##### Research question R2

What socioeconomic and demographic factors are associated with patients visiting a public or a private healthcare facility to seek care for illness conditions of perceived mild severity?

The null and alternate hypotheses associated with the above research question are given below.

H_0_: Socioeconomic and demographic attributes do not influence a patient’s choice of visiting a public or a private healthcare facility in seeking care for illness conditions of perceived mild severity.

H_1_: Socioeconomic and demographic attributes influence a patient’s choice of visiting a public or a private healthcare facility in seeking care for illness conditions of perceived mild severity.

The BLR model for *R2*, with significant variables alone, is given below.

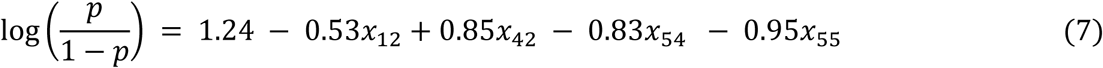

p = probability of choosing a public healthcare facility

*x*_12_ = 1, if the respondent is female

*x*_42_ = 1, if the respondent has completed 10^th^ grade

*x*_54_ = 1, if the respondent has annual income between USD 23,000 – USD 46,000

*x*_55_ = 1, if the respondent has annual income above USD 46,000

We see from Table 5 that patient gender had a significant association with patient HSB, with female patients preferring to visit a private healthcare facility 1.72 times more than a public facility in case of illness conditions of perceived mild severity. There appears to be evidence from other parts of the world that corroborates our findings. Keesara et al. (2015) studied preferences of women for public and private healthcare providers in Kenya and reported that a significant proportion of women preferred visiting private healthcare facilities due to convenience and timeliness in receiving care, and also were willing to pay more for private care. This study focused specifically on women visiting health facilities for family planning. This observation was also conveyed to us by a few male patients, who indicated that their spouses preferred visiting private healthcare facilities even in cases of mild ailments. Other patient attributes significantly affecting HSB for illness conditions of perceived mild severity were education and annual income.

##### Research question R3

What socioeconomic and demographic factors are associated with patients visiting a public or a private healthcare facility in the case of illness conditions of perceived high severity?

The null and alternate hypotheses associated with *R3* are given below.

H_0_: Socioeconomic and demographic attributes do not influence a patient’s choice of visiting a public or a private healthcare facility in seeking care for illness conditions of perceived high severity.

H_1_: Socioeconomic and demographic attributes influence a patient’s choice of visiting a public or a private healthcare facility in seeking care for illness conditions of perceived high severity.

We find, as shown in Table 5, that employment status, annual level of income, and number of children were notable factors influencing patient HSB for illness conditions of perceived high severity. From equation 8 below, we see that employed patients with higher levels of income were more likely to visit a private healthcare facility for getting treatment for their medical condition. The odds of visiting a private healthcare facility increased by 2 times and 3.03 times among patients with a source of employment and having an annual income greater than USD 46,000 in comparison to students and patients without any source of income, respectively. Patients having two or more than two children were 3.16 times and 3.05 times more likely to visit public healthcare facilities in comparison to patients without children.

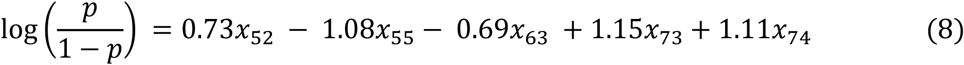

p = probability of choosing a public healthcare facility

*x*_52_ = 1, if the respondent has annual income is between USD 11,500 – USD 23,000

*x*_55_ = 1, if the respondent has annual income greater than USD 46,000

*x*_63_ = 1, if the respondent is employed

*x*_73_ = 1, if the respondent has 2 children

*x*_74_ = 1, if the respondent has more than 2 children

##### Research question (R4)

What socioeconomic and demographic factors are associated with a patient visiting a primary or a specialized healthcare facility for their first visit upon falling ill?

*For* R4, of the two complementary categories for this response variable - (a) primary healthcare facilities, and (b) specialized healthcare facilities - we chose the latter as the reference category. The null and alternate research hypotheses are given below.

H_0_: Socioeconomic and demographic attributes do not influence a patient’s choice of visiting a primary or a specialized healthcare facility for their first visit.

H_1_: Socioeconomic and demographic attributes influence a patient’s choice of visiting primary or a specialized healthcare facility for their first visit.

The BLR model for *R4*, with significant variables alone, is expressed as:

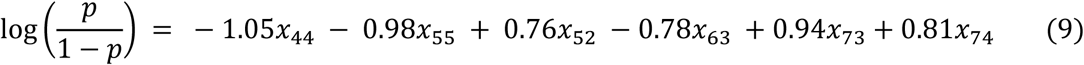

p = probability of choosing a primary level healthcare facility

*x*_44_ = 1, if the respondent has undergraduate degree

*x*_45_ = 1, if the respondent has post-graduate degree

*x*_52_ = 1, if the respondent has annual income between USD 11,500 – USD 23,000

*x* _63_ = 1, if the respondent is employed

*x* _73_ = 1, if the respondent has 2 children

*x*_74_ = 1, if the respondent has more than 2 children

We report parameter estimates of significant variables for *R4* in Table 5. Higher education levels along with higher annual incomes were associated with increased odds of visiting specialized healthcare facilities such as clinics and hospitals. Patients with undergraduate and post-graduate degrees were 2.94 times and 2.70 times more likely to visit specialized facilities directly in comparison to patients without any formal education.

#### 3.2.2 MLR based HSB model

We examined the socioeconomic and demographic factors associated with patient preferences for seeking care – upon first falling ill - from different types of medical practitioners such as general physicians, specialized doctors, doctors formally trained in traditional medicine, and informal traditional healers. As there were more than two categories for the response variable, we implemented the MLR modelling technique.

##### Research question (R5)

What factors are associated with patient preferences for visiting different types of medical practitioners - i.e., general physicians, specialized doctors, doctors formally trained in traditional medicine, or informal traditional healers – upon first falling ill?

The null and alternate hypotheses for *R5* are given below.

H_0_: Socioeconomic and demographic attributes of patients are not associated with their choice of medical practitioner type.

H_1_: Socioeconomic and demographic attributes of patients are associated with their choice of medical practitioner type.

The predictor variables are described in Figure 2, and we chose ‘general physician’ as the reference category and built three individual BLR models with respect to the reference category. Six independent variables - marital status, age, education level, annual income level, number of children, and employment status - turned out to be significant predictors. We report the parameters of significant independent variables for all three models in separate rows in Table 6.

The MLR models, with significant variables alone, are provided below.

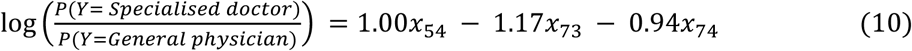

*x*_54_ = 1, if the respondent’s annual income is between USD 23,000 – USD 46,000

*x*_73_ = 1, if the respondent has two children

*x*_74_ = 1, if the respondent has three or more than three children

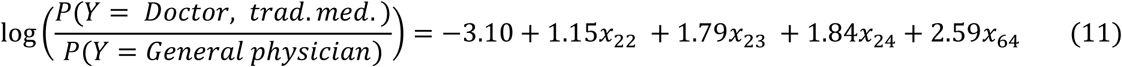

*x*_22_ = 1, if the respondent is between 30-39 years of age

*x*_23_ = 1, if the respondent is between 40-49 years of age

*x*_24_ = 1, if the respondent is between 50-59 years of age

*x*_64_ = 1, if the respondent’s is unemployed

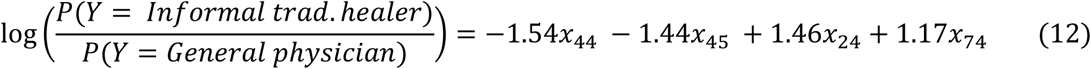

*x*_24_ = 1, if the respondent is between 50-59 years of age

*x*_44_ = 1 if the respondent has undergraduate degree

*x*_45_ = 1 if the respondent has post-graduate degree

*x*_74_ = 1 if the respondent is unemployed

We made the following inferences from equations 10, 11, and 12.

1. Patients with higher annual income levels preferred directly consulting specialized practitioners. Specifically, patients with income level between USD 23,000 – 46,000 were 2.71 times more likely to visit specialized doctors in comparison with patients with annual income level up to USD 11,500.
2. Patients with two or more than children were 3.22 times and 2.56 times more likely to visit a general physician in comparison to patients without any children.
3. Patients aged between 30 – 39 years, 40 – 49 years, and 50 – 59 years consulted doctors formally trained in traditional medicines 3.15 times, 5.98 times, and 6.29 times respectively more than patients aged between 18 – 29 years.
4. Unemployed patients were 3.22 times more likely to visit traditional healers in comparison to students.
5. We observed that patients with undergraduate and/or post-graduate degrees were 0.18 times and 0.21 times less likely to visit a traditional healer in comparison to patients without any formal education. Previous studies conducted in the other countries discussed how higher level of education raised awareness among patients and further increased the likelihood of seeking formal care (Hahn & Truman, 2015; Zajacova & Lawrence, 2018).

**Table 8:**
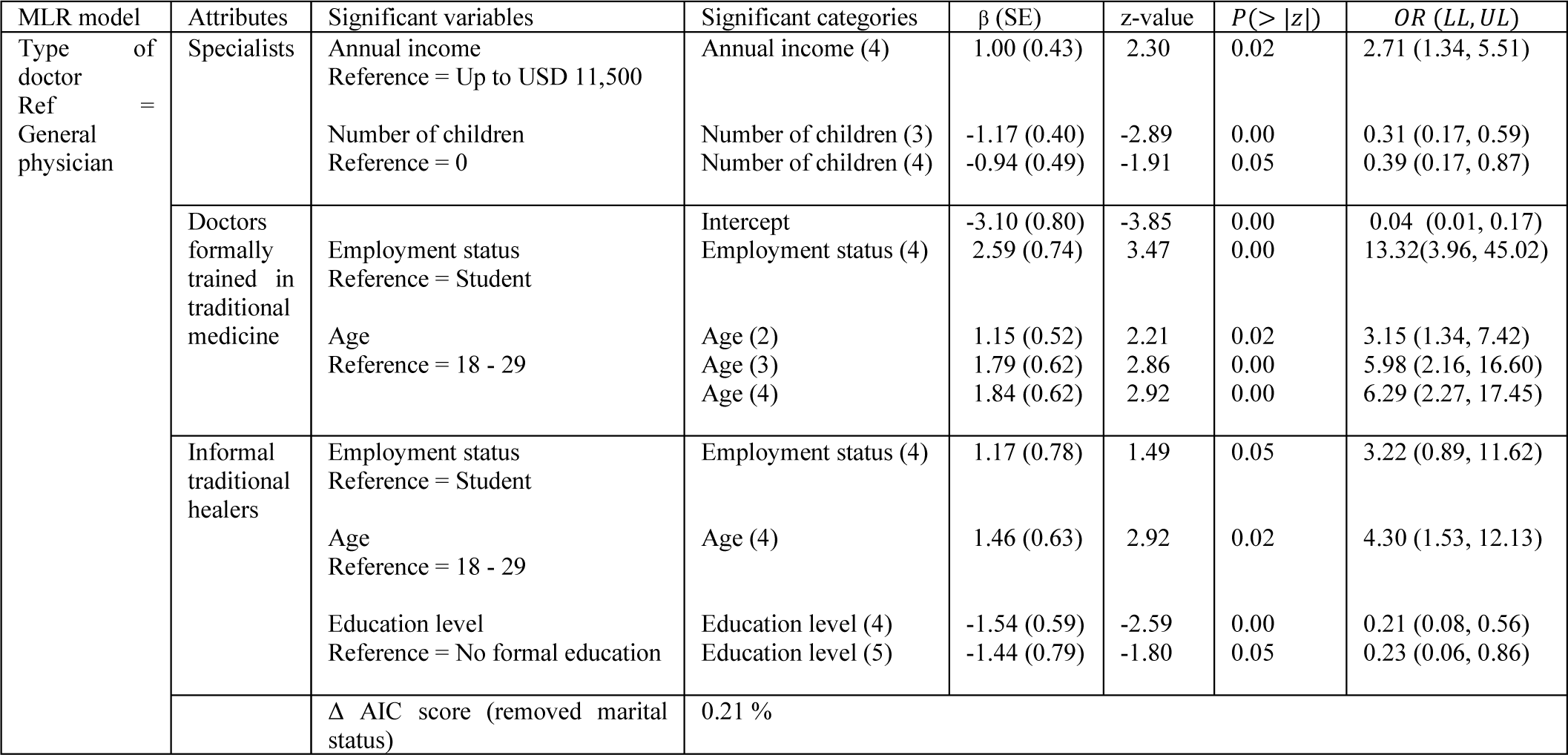
Results from MLR modelling of HSB as a function of socioeconomic and demographic variables.

## 4. Discussion

In this study, we examined the HSB of patients visiting primary and secondary public healthcare facilities in an urban Indian district. Based on our survey responses, we used logistic regression to model the association between various aspects of patient HSB and their socioeconomic and demographic attributes. Our analyses provided quantitative evidence for the association of HSB with socioeconomic and demographic factors such as annual income, education level, occupation, gender, age and other factors such as perception of expected wait time and cleanliness at healthcare facilities, expected quality of care, and behavior of service providers.

We now discuss specific findings of our work. We observed that a significant proportion of patients (40.44%) preferred to make a first visit to specialized healthcare facilities (public or private) for treatments. This finding is consistent with the work by Narang (2010), and Rao & Sheffel (2018) who find that a majority of patients bypassed primary and secondary care facilities to directly seek care from specialists at higher level facilities. This practice adversely affects the hierarchy in patient flow across the public healthcare facility network, and can lead to both overcrowding at facilities offering a higher level of care and underutilization of lower-level facilities. Thus our findings for an urban Indian district, in conjunction with the findings by Rao & Sheffel (2018) for primary healthcare centres (PHCs) in a rural Indian region, emphasize the need for a comprehensive pan-Indian investigation into the reasons for bypassing public primary and secondary healthcare facilities. Rao & Sheffel (2018) discuss that improving structural quality of PHCs alone is unlikely to suffice, and that improvement in provider attitudes and the quality and quantity of time spent with patients is likely to be required. Our findings also support this: we find that patients choose to visit facilities where they have had prior good experiences not only in terms of adequate infrastructure, but also in terms of quality of care, and provider attitudes. However, improving quality of care at primary and secondary care facilities may also need to be accompanied by the implementation and enforcement of an effective referral mechanism across the public healthcare network to alleviate the problem of overcrowding at higher levels of care. Similar mechanisms exist in developed nations such as Britain, France, Germany, Singapore, South Korea, etc., where deviating from predefined referral pathways may lead to penalties for non-urgent cases in terms of delayed reimbursement, higher copayment, or longer wait times (P. Jia et al. 2017; You et al. 2019).

We also observed that a significant proportion of patients (80.14%) preferred visiting private healthcare facilities which charge significantly higher medical expenses over visiting public facilities for specialized medical services required to treat serious and/or chronic illnesses. Similarly, patients with higher incomes were 4.34 times more likely to visit private healthcare facilities in comparison to patients with lower incomes. This might be due to excessive waiting times at public facilities prior to receiving care, unavailability of specialized medical equipment, inadequate numbers of inpatient beds, insufficient time spent with patients by providers, or unhygienic conditions. Given India’s large population, it may not be feasible for public healthcare facilities alone to cater to the healthcare needs of the entire populace. However, our study illustrates that an investigation into factors that discourage patients (in particular, those with low annual incomes) from seeking care at public healthcare facilities, especially for illnesses of perceived high severity, is warranted.

From the perspective of gender, female patients also indicated a higher preference for private healthcare providers. This is consistent with findings from Kenya, where Keesara et al. (2015) also reported that women avoided public facilities owing to long waiting times and impolite care providers. This suggests that public health planning authorities may need to identify and implement ways to build and improve trust among female patients for public facilities. In addition, patients with lesser education and without any source of income preferred visiting traditional healers or took medicines without consulting a formal care provider. This may indicate the need for targeted programs to improve awareness among these groups regarding the provision of free or nominally priced care at public facilities.

A key limitation of this work is that the HSB analyses are based on the survey conducted in the South-west Delhi district alone. Thus the findings may not be representative of the entire population of New Delhi or other urban regions. For obtaining a more comprehensive picture of HSB in India, similar surveys may be conducted in other regions of India as well. Our study provides a template for conducting such studies, analyses that can be done from the data collected via these studies, and the insights that can be generated – not only for India, but also for other developing nations with a similarly complex healthcare landscape.

## Data Availability

Survey data can be shared upon request.

## Acknowledgement

This work was supported by the Prime Minister’s Research Fellowship (PMRF), Government of India.

## Notes

### Competing Interest Statement

The authors have declared no competing interest.

### Funding Statement

Najiya Fatma is funded by the Prime Minister's Research Fellowship programme funded by the Government of India.

### Author Declarations

The Institutional Review Board at the Indian Institute of Technology New Delhi approved the study protocol with approval number IITD-IEC-ID-P064 on November 25, 2019.

